# Cross sectional study on behavioral determinants associated with sugary foods and beverages and its corelates with body mass index among adolescents

**DOI:** 10.1101/2024.03.24.24304815

**Authors:** Laxmi Gautam, Milan Thapa, Poonam Pokhrel, Parash Mani Sapkota, Anjali Bhatt, Kiran Paudel

## Abstract

**Background:** Adolescence is the period that demands high nutrients with changes in dietary habits making them vulnerable. Foods high in calories and deficit in essential nutrients increases risk for overweight. The habits are guided by multiple factors including behavioral determinants. Therefore, this study aims to identify the status of and factors associated with behavioral determinants on adolescents in regard to sugary items.

**Methods:** A cross-sectional study was conducted among 768 adolescents of Nagarjun Municipality, Kathmandu from September 2022 to January 2023. This study used a multistage random sampling. The chi square test and logistic regression were applied to analyze the results in SPSS V.16 and p-value ≤ 0.05 was considered as statistically significance.

**Result:** Adequate level of knowledge was found among 84.11% (95% CI: 81.52 to 86.70) of the adolescents. Positive attitude was seen among 60% of the adolescents (95% CI: 56.55 to 63.49). The percentage of adolescent consuming items was 84.50% (95% CI: 81.94 to 87.07). Odds of having adequate knowledge among respondents was twice (AOR=2.05, 95% CI: 1.12 to 3.76) more likely for those who were living with their parents. Female adolescents (AOR=2.51, 95% CI: 1.61 to 3.89), whose mother are homemaker (AOR=1.99, 95% CI: 1.04 to 3.58) and father are engaged in foreign employment (AOR = 2.09, 95% CI: 1.04 to 4.21) were more likely to consume sugary items. Prevalence of overweight was seen among 6.38% (95% CI: 4.64 to 8.11) of respondents. Consumption was seen to be significant to the model overweight versus normal [OR=11.95 (95% CI: 1.61 to 88.42)].

**Conclusion:** Presence of adequate knowledge wasn’t the only and adequate factor for food selection. Family indulged interventions can be useful as familial factors seem to be affecting behavioral characteristics. Sugary foods and beverages are associated with overweight. Hence interventions focusing on dietary discipline is also recommended.

**STRENGTHS AND LIMITATIONS OF THE STUDY:**

- This study deviates the primary focus from range of processed foods items to sugary foods and beverages only. No such studies with sole focus on high calorie sugary foods have been found conducted in Nepal.
- Study was limited to a single municipality, whose results cannot be generalized for national context. However, it can apply in case of urban settings.
- Majority of the findings are based on self-reporting, which leaves enough space for under-reporting and recall bias. On the other hand, anthropometric measurements taken by research team was positive part of the study.
- Difficult to gauge the impact of the findings, because weight gain in adolescence phase varies as a function of age, maturation, and growth velocity not just consumption of sugary foods and beverages.

## INTRODUCTION

Added sugars are not an essential part of a healthy diet, despite they give sensory effects to the meals and increase enjoyment. They can replace nutrient-dense meals and contribute to poor health outcomes by supplying calories without delivering other necessary nutrients, which is especially concerning in youngsters including adolescents.^1^ Consuming sugar in excess has been associated with a number of metabolic irregularities and harmful health effects including high risks of Non-Communicable Diseases (NCDs) and even creates nutrition insufficiency in individual’s body.^2^ Unhealthy diets are one of the most decorated and heavily linked modifiable risk factors for NCDs.^3^

Evidence from past and recent studies shows that the lifestyle, dietary habits, and behavioral patterns formed during adolescence can remain into adulthood and have a substantial impact on one’s health.^4,5^ Childhood and adolescent obesity raises the chance that an adult may continue to weigh more than what is healthy in latter phases of life as well.^4,5^ Excessive consumption of sugar in various forms most especially the sugary food items and beverages has been one of the crucial factor for promoting overweight and obesity all around, including Lower Middle Income Countries like Nepal.^6,7^ Adolescence is the phase of life between childhood and adulthood, from ages 10 to 19. It is a unique stage of human development and an important time for laying the foundations of good health.^8^ The nutrition and food habits during the phase is extremely important and determines the health in adult stage of life and bad dietary and eating habits are the most prevalent risk factor for chronic illness in children and adolescents aged 12 to 17 years.^9^ Childhood exposure to sugary foods and beverages can influence an adult’s liking for sweet meals so an early-life nutrition is crucial for a child’s development as well as being a modifiable risk factor for non-communicable diseases (NCDs).^10–12^ Since adolescence is a period that demands high nutrients with changes in dietary habits among adolescents, it makes the age group more favorites for getting exposed in consumption of heavily processed and high sugar products.^13–15^

Ultra-processed foods and non-alcoholic beverages with high amount of trans-fat, free sugar or salt are being heavily consumed by millions of children and adolescents worldwide which is ultimately bringing harmful effects for the health and development of those population.^16–18^ Energy dense foods and sugary drinks contribute to weight gain, overweight and obesity by promoting excess energy intake.^6^ Recent figures generated from several studies also showed that the prevalence of overweight has tripled in many countries, making it the major public health issue in the 21st century.^19^ Sugar sweetened beverages have been found as a unique dietary contributor to obesity.^20^ Six percentage of children and adolescents aged 5-19 are expected to be obese by 2030, according to the projections from NCD risk factor collaboration.^21^ It was reported that 33.3% of adolescents had habit of drinking carbonated drinks according to a survey conducted in 2015.^22^ The processed food consumption rate among the kids especially from Kathmandu valley is abruptly increasing.^23^ Multiple factors including personal traits, physical and societal spheres affect the dietary choice including knowledge, attitude and beliefs which ultimately guides the practice. This study aims to assess and understand the awareness level and consumption patterns of sugary foods and beverages among adolescents.

## METHODS

### Study area

The study was conducted in ten schools (5 private and 5 public) which were selected randomly from the pool of 58 schools of Nagarjun Municipality, Kathmandu district. Nagarjun Municipality is a fast-urbanizing local level located in northwest of Kathmandu valley and the second most populated administrative unit of Kathmandu district. The total adolescent population in the local level is 18647^24^ which makes up 16.15% of the total population.

### Study design

A cross sectional study was conducted among the school going adolescents. The study was conducted from August 2022 to February 2023.

### Sampling procedure

A multistage random sampling method was used to select the participants. Initially five random wards were selected from the total of ten wards inside the Nagarjun Municipality. There were 58 schools (13 public and 45 private) with a total of 4191 students in the sampling frame inside those five wards. A complete list of all schools inside Nagarjun Municipality was obtained from education section of the municipality itself. Schools with less than total of 50 students combined in grade eight, nine and ten were excluded from the study. Eventually the remaining 34 schools were included; 10 public school and 24 private school with 3829 students. Finally, 2 schools (one public and one private) were randomly chosen from each of the five wards selected. Then, all the students were selected from those same ten schools. Primary schools and adolescents who were absent on the days of data collection were excluded from the study.

### Sample size determination

The sample size was calculated using single population proportion formula, 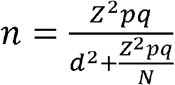 considering 95% confidence interval (CI), 50% assumed proportion^25^ (p), and 5% margin of error (d). Assuming a 10% non-response rate and 2 design effect, the sample size was estimated to be 768.

### Questionnaire design

A semi-structured questionnaire was developed for the study based on study objectives by the research team after vigorous literature review and consultation with faculty members of public health who’d been involved in research for multiple years. The questionnaire basically comprised of various sections comprised of socio-demographic characteristics, parental characteristics, questions to assess knowledge regarding sugary foods and beverages and related health impacts (9 questions), questions to assess attitude towards sugary foods and beverages (13 questions/ statements) and questions to assess the consumption of sugary foods and beverages (9 questions). Evaluation of knowledge and practice was done by assigning ‘1’ for positive/correct answer/statement and ‘0’ for negative/incorrect answer/statement. However, eleven items related to the sugary foods and beverages alongside ill-effects were used to access attitude. The responses based on Likert’s scaling technique had three possible levels, ranging from agree to disagree. The evaluation of attitude was done by assigning ‘2’ for positive/correct answer/statement, ‘1’ for neutral stand and ‘0’ for negative/incorrect answer/statement. Participants that scored the median of the total score or above were considered having an adequate knowledge and positive attitude. A score of 5 or above was considered as an adequate level of knowledge while respondents’ scores of 19 or above were considered good attitude levels based on the total attainable score of 22, referenced to the median score attained from Shapiro-Wilk test (p<0.05). A pilot study of the questionnaire was performed in one non-sampled school which was found to be appropriate. A copy of the study questionnaire can be found in online Supplementary File 1.

### Data collection and statistical analysis

Self-administered questionnaires were used for data collection from September 2022 to October 2022. Anthropometric measurements (height and weight) were obtained using a calibrated digital bathroom weighing scales and a standard tape measure. The accuracy of the instruments was checked using the standard weight and height at the beginning of every data collection session. Statistical analysis was performed using SPSS V.16. Association between independent variables and the dependent (Knowledge, Attitude, Practice) and outcome variables (Body Mass Index) was assessed through Chi-square test and logistic regression analysis. The Hosmer–Lemeshow test was performed to test the goodness-of-fit of the multivariate logistic regression model, and the model was found to be a good fit (p>0.05).

### Ethical consideration

All the study related activities were initiated after obtaining the relevant approval from the Institutional Review Committee of Manmohan Memorial Institute of Health Sciences (MMIHS IRC 887; Ref 79/151). Prior permission and written consent were also taken before the data collection from education section of the municipality and school administration. Participants were also clearly informed regarding the study before the start. Confidentiality of information was assured and ensured throughout the study. The Strengthening the Reporting of Observation Studies in Epidemiology (STROBE) cross sectional reporting guidelines^26^ was also used to ensure the quality of reporting and transparency.

## RESULTS

General demographic characteristics of the study participants is presented in Table 1. The median (±IQR) age of adolescents was 15±1 years, ranging between 11 and 18 years of age. Adolescents under fourteen years old comprised 52.5%, slightly more than those aged ≤ 14 years (47.5%). There were more male adolescents (51.4%) than females. Half of the respondents belonged to the Janajati group (50.1%), while 44.4% were from the Brahmin/Chhetri ethnic group. The majority (75.8%) followed the Hindu religion. When asked about their living situation, 91.1% of respondents lived with their parents, while the rest resided with relatives, friends, or alone. Concerning household income, most of the parents (33.8%) earned between NPR 15,001-30,000 per month. Over one-third of the mothers (34.4%) had attained secondary level of education and 41.6% were homemakers. Similarly, higher share of fathers had also attained secondary level education (41.1%), while nearly a third (32.9%) were employed in job services, making it the highest form of professional engagement among fathers.

**Table 1.**
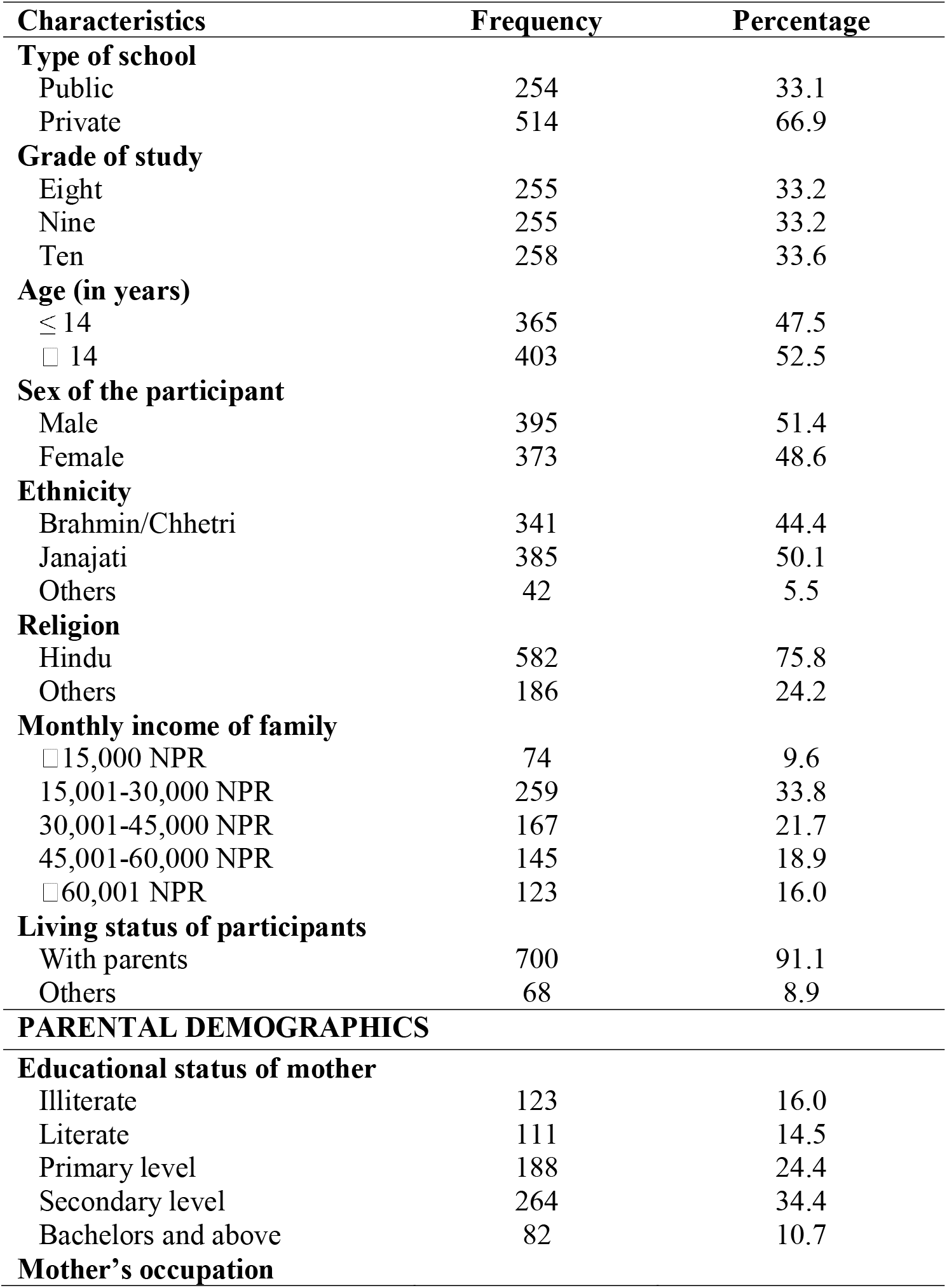
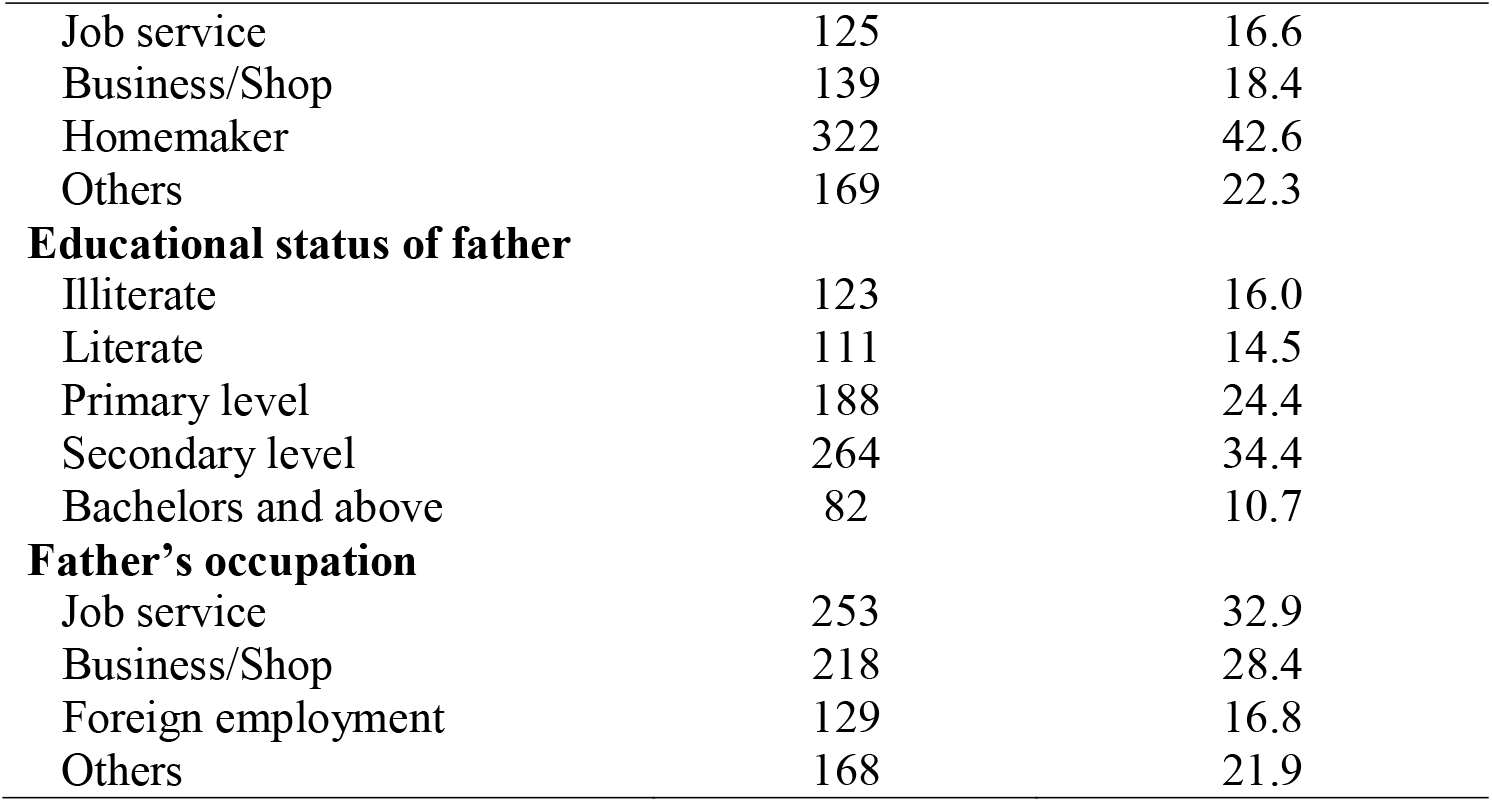
Socio-demographic characteristics of study participants.

| **Characteristics** | **Frequency** | **Percentage** |
| --- | --- | --- |
| **Type of school** |  |  |
| Public | 254 | 33.1 |
| Private | 514 | 66.9 |
| **Grade of study** |  |  |
| Eight | 255 | 33.2 |
| Nine | 255 | 33.2 |
| Ten | 258 | 33.6 |
| **Age (in years)** |  |  |
| ≤ 14 | 365 | 47.5 |
| > 14 | 403 | 52.5 |
| **Sex of the participant** |  |  |
| Male | 395 | 51.4 |
| Female | 373 | 48.6 |
| **Ethnicity** |  |  |
| Brahmin/Chhetri | 341 | 44.4 |
| Janajati | 385 | 50.1 |
| Others | 42 | 5.5 |
| **Religion** |  |  |
| Hindu | 582 | 75.8 |
| Others | 186 | 24.2 |
| **Monthly income of family** |  |  |
| ≤ 15,000 NPR | 74 | 9.6 |
| 15,001-30,000 NPR | 259 | 33.8 |
| 30,001-45,000 NPR | 167 | 21.7 |
| 45,001-60,000 NPR | 145 | 18.9 |
| > 60,001 NPR | 123 | 16.0 |
| **Living status of participants** |  |  |
| With parents | 700 | 91.1 |
| Others | 68 | 8.9 |
| **PARENTAL DEMOGRAPHICS** |  |  |
| **Educational status of mother** |  |  |
| Illiterate | 123 | 16.0 |
| Literate | 111 | 14.5 |
| Primary level | 188 | 24.4 |
| Secondary level | 264 | 34.4 |
| Bachelors and above | 82 | 10.7 |
| **Mother's occupation** |  |  |
| Job service | 125 | 16.6 |
| Business/Shop | 139 | 18.4 |
| Homemaker | 322 | 42.6 |
| Others | 169 | 22.3 |
| **Educational status of father** |  |  |
| Illiterate | 123 | 16.0 |
| Literate | 111 | 14.5 |
| Primary level | 188 | 24.4 |
| Secondary level | 264 | 34.4 |
| Bachelors and above | 82 | 10.7 |
| **Father's occupation** |  |  |
| Job service | 253 | 32.9 |
| Business/Shop | 218 | 28.4 |
| Foreign employment | 129 | 16.8 |
| Others | 168 | 21.9 |

More than four-fifths (81.4%) of the respondents considered sugary foods and beverages foods to be harmful for health. The prevalence of adequate knowledge regarding sugary foods and beverages and ill health effects caused due to them among adolescents was 84.11% (95% CI: 81.52-86.70) as shown in Table A of Supplementary File 2. Exactly three fifths (95% CI: 56.55-63.49) of the respondents held a positive attitude (means they were aware and positive towards the harmfulness and ill effects of sugary foods and beverages). The rate of consumption of sugary foods and beverages among the adolescents was 84.50% (95% CI: 81.94-87.07). Of all respondents consuming sugary foods and beverages, highest (68.1%) share of respondents consumed confectionaries and bakeries like ice-cream, ice pops, cake, pastries followed by cold sugary beverages (66.4%) like Sugar Sweetened Beverages, carbonated/aerated drinks, artificial fruit juice and energy drinks as shown in Table C of Supplementary File 2. More than four-fifths (81.8%) of respondents felt the need to control and limit the intake of sugary foods and beverages, while just slightly more than three-fifth (62.5%) of them were confident about discontinuing the consumption if necessary. (Figure 1)

**Figure 1.**
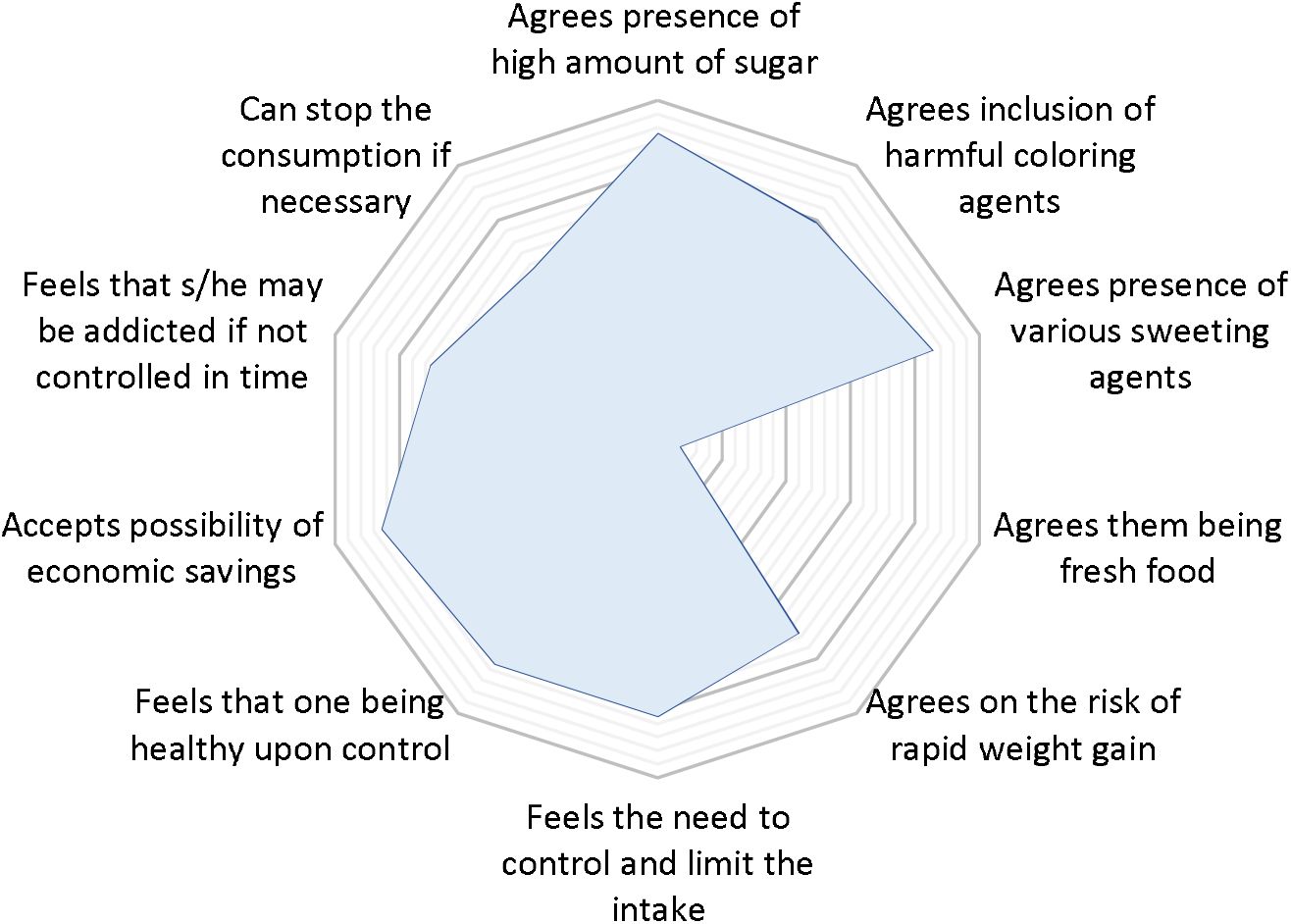
Attitude towards sugary foods and beverages alongside its effects

What stands out in the Table H of Supplementary File 2 is the general pattern of Body Mass Index (BMI) seen among the adolescents during the study. It was revealed that out of 768 adolescents, half (50.4%) of the participants had weight in normal range. Similarly, 6.38% (95% CI: 4.64-8.11) of the respondents were found to have high BMI considerably categorized into overweight and obese.

Table 2 represents bivariate and multivariate logistic regression between behavioral determinants and socio-demographic characteristics. Odds of having adequate knowledge among respondents was twice (AOR=2.05, 95% CI: 1.12-3.76) more likely for those who were living with their parents as compared to the other counterpart adjusting other explanatory variables. Female adolescents were 2.51 times (AOR=2.51, 95% CI: 1.61-3.89) more likely to consume sugary foods and beverages as compared to their counterpart male adolescents adjusting remaining predictors. Adolescents whose mother were homemaker have an odd of consuming sugary foods and beverages 1.82 times (AOR=1.99, 95% CI: 1.04-3.58) more than the respondents whose mothers are involved in job services. Similarly, adolescents whose fathers were aboard for foreign employment were 2.09 times (AOR = 2.09, 95% CI: 1.04–4.21) more likely to consume sugary foods and beverages as compared to adolescents whose father were involved in job services.

**Table 2.** Factors affecting knowledge and practice regarding sugary food and beverages among adolescents

| Demographic Characteristics | Knowledge |  | Practice |  |
| --- | --- | --- | --- | --- |
|  | COR (95% CI) | AOR (95% CI) | COR (95% CI) | AOR (95% CI) |
| **Type of school** |  |  |  |  |
| Public | Ref | Ref | Ref | Ref |
| Private | **1.84 (1.24-2.73)\*** | 1.45 (0.90-2.34) | **1.51 (1.01-2.26)\*** | 1.49 (0.93-2.38) |
| **Grade of study** |  |  |  |  |
| Eight |  |  | Ref |  |
| Nine |  |  | 1.64 (0.98-2.74) |  |
| Ten |  |  | 0.88 (0.56-1.39) |  |
| **Sex of participant** |  |  |  |  |
| Male |  |  | Ref | Ref |
| Female |  |  | **2.37(1.56-3.61)\*** | **2.51 (1.61-3.89)\*** |
| **Monthly income of family** |  |  |  |  |
| □ 15,000 | Ref | Ref |  |  |
| 15,001-30,000 | 1.76 (0.96-3.22) | 1.32 (0.68-2.56) |  |  |
| 30,001-45,000 | **2.31 (1.18-4.55)\*** | 1.36 (0.62-3.00) |  |  |
| 45,001-60,000 | **2.45 (1.21-4.96)\*** | 1.30 (0.56-3.01) |  |  |
| □ 60,001 | **2.66 (1.26-5.61)\*** | 1.23 (0.50-3.04) |  |  |
| **Living status of adolescents** |  |  |  |  |
| With parents | **2.06 (1.15-3.67)\*** | **2.05 (1.12-3.76)\*** |  |  |
| Others (Relatives/Hostel/Alone) | Ref | Ref |  |  |
| **Mother's occupation** |  |  |  |  |
| Job service |  |  | Ref | Ref |
| Business/Shop |  |  | 1.31 (0.70-2.43) | 1.13 (0.61-2.09) |
| Homemaker |  |  | **2.08 (1.20-3.62)\*** | **1.99 (1.10-3.58)\*** |
| Others |  |  | 1.05 (0.59-1.86) | 1.07 (0.56-2.03) |
| **Father's occupation** |  |  |  |  |
| Job service | Ref | Ref | Ref | Ref |
| Business/Shop | 1.06 (0.63-1.78) | 0.94 (0.55-1.60) | 1.71 (0.71-1.90) | 1.14 (0.67-1.94) |
| Foreign employment | 1.34 (0.70-2.54) | 1.51 (0.77-2.94) | **1.93 (1.00-3.72)\*** | **2.09 (1.04-4.21)\*** |
| Others | **0.60 (0.36-0.99)\*** | 0.75 (0.43-1.29) | 1.13 (0.67-1.90) | 1.22 (0.68-2.19) |

Multinomial logistic regression between behavioral determinants and BMI has been shown in Table 3. Body Mass Index of adolescents with three categories-underweight, normal and overweight/obesity was taken as the dependent variable and behavioral determinants; knowledge, attitude and practice as predictors/regressors. Consumption of sugary foods and beverages was found to be significant to both the models of normal versus underweight and normal versus overweight/obesity. The odds ratio of consumption of sugary foods and beverages as compared to no consumption is 1.58 (95% CI: 1.05-2.39) and 11.95 (95% CI: 1.61-88.42) for underweight relative to normal and overweight/obese relative to normal BMI, respectively. This indicates that the adolescents who consume sugary foods and drinks are 1.58 times and 11.95 times more likely to have underweight relative to normal BMI and overweight/obese to normal BMI respectively, compared to adolescents who do not consume sugary foods and beverages. The coefficient is positive for both the model with significant value suggesting that higher consumption increases the odds of underweight and overweight.

**Table 3.** Influence of behavioral determinants on Body Mass Index (BMI)

| Behavioral determinants | Model 1 |  | Model 2 |  |
| --- | --- | --- | --- | --- |
| | Estimated coefficient | Odds Ratio ($e^{\beta}$) (95% CI) | Estimated coefficient | Odds Ratio ($e^{\beta}$) (95% CI) |
| Knowledge regarding sugary foods and beverages |  |  |  |  |
| Inadequate knowledge |  | Ref |  | Ref |
| Adequate knowledge | -0.32 | 0.72 (0.47-1.09) | -3.792 | 0.55 (0.25-1.20) |
| Attitude towards sugary foods and beverages |  |  |  |  |
| Negative attitude |  | Ref |  | Ref |
| Positive attitude | -0.27 | 0.75 (0.55-1.02) | -0.583 | 0.88 (0.47-1.65) |
| Practice of sugary foods and beverages |  |  |  |  |
| Do not consume |  | Ref |  | Ref |
| Consume | 0.46 | **1.58 (1.05-2.39)\*** | 2.481 | **11.95 (1.61-88.42)\*** |
| **Model 1:** Normal vs Underweight |  |  |  |  |
| **Model 2:** Normal vs Overweight/Obesity |  |  |  |  |
| **\*p value <0.05** |  |  |  |  |

## DISCUSSION

Various studies have been conducted to examine behavioral determinants on processed foods previously; however, most studies have not exclusively focused on sugar-rich foods and beverages in specific to the best of our knowledge. Our study suggests adolescents living with their parents had higher odds of having adequate knowledge regarding sugary foods and beverages and ill-effects associated with them. Similarly, being female, children of homemaker mother and father employed in foreign had higher odds of consuming sugary foods and beverages. The odds of being overweight/obese was higher among the adolescents consuming sugary foods and beverages.

Adequate level of knowledge regarding sugary foods and beverages and harmful impacts associated was found among 84.1% of the study participants in this study which is more that the figure found in a study conducted among adolescents of Pokhara valley where only two-third (66.5%) of the adolescents had adequate knowledge regarding harmful effects of processed foods.^27^ The yields in this study were higher compared to those of other studies conducted by Upreti et al. and Subedi et al. in Chitwan where the adequate knowledge among adolescents were found to be 55.9% and 65.6% respectively.^28,29^ However, this finding accord with another study done by Paudel et al.^30^ indicating higher number of respondents with adequate knowledge regarding health effects of processed foods including sugary items among comparable population which is obvious due to similarity in population and study settings. The finding is contrary to previous studies conducted in surrounding neighboring nations including India^31^, Vietnam and Thailand^32^ with similar context where low level of adequate knowledge was seen among adolescents regarding processed foods and sugary beverages. A possible explanation for these results may be the lack of adequate curriculum for adolescents focused on nutrition and related issues among the referenced nations.

Knowledge level was not found to be associated with the age of respondents alike an Indian study conducted by Joseph et al.^33^ Odds of having adequate knowledge among respondents was twice more likely for those who were living with their parents as compared to the other counterpart adjusting other explanatory variables in our study. Similar findings were seen in another study conducted in Kathmandu.^34^ This might be due to the reason that good family environment and parental guidance shapes the food selection process with adequate access to information regarding the issues which is also further supported by the fact that around four-fifth of the parents were found to pressurize their children against consumption of sugary foods and beverages in this study. Three fifths (60.0%) of the respondents held a positive attitude on the issue of the study. The yields in this investigation were higher compared to those of other studies, where the attitude of respondents was inclined unlike this study. A cross sectional study conducted in Manglore city of India, reported positive attitude among slightly less than half (48.3%) of the participants.^33^ The level observed in this study were far below than those observed by Upreti et al. among adolescents (88.4%) in Chitwan.^28^ Similarly, findings from a similar study on sugar sweetened beverages conducted by Thanh Ha et al. showed more than two thirds (68.5%) of the respondents had positive attitude, even more than that of this study.^35^

The overall level of consumption of sugary foods and beverages in the study was 84.5 percent, which is higher than those found in other studies, where confectionaries and sugary beverages was consumed by 58% and 48% of the respondents respectively.^27^ However into the categorization, this study also yielded 69% and 68% in those respective food groups, which is still a bit higher. A relatively lower frequency was observed in chocolate selection in a study conducted among adolescents in India^33^ and also a slightly higher age group in Nepal.^34^ It is possible that these results merely reflect a selection effect, as the referenced study undertook only male particiants^33^ and higher age group^34^ who may have opted for other selection in processed foods rather than sweets. However these results reflect those of Paudel et al. who also founded higher proportion of study participants consuming chocolates (72.6%) and cold drinks (52%) during the study.^30^ A large proportion of adolescents were found to be consuming sugary beverages in Malaysia.^36^

Consumption was seen to be associated with gender of adolescent, occupation of mother and father. Knowledge level was not found to be associated with consumption status alike some previous studies.^30,33^ No associations were seen between the socio-demographic characteristics and consumption status of Sugar Sweetened Beverages in a study conducted among school adolescents in Selangor, Malaysia.^36^ Around one third of the respondent were consuming sugary foods and drinks due to influence of good taste in this study. Similar findings had been seen in other studies as well where highest (52%) number of the adolescents consuming processed foods mentioned taste as the major reason for consumption in a study conducted by Subedi et al among secondary level students in Ratnanagar Municipality.^29^ Vast majority (91%) of the adolescents were also consuming processed foods due to taste in another study conducted among adolescents of Kathmandu valley by Paudel et al.^30^ Large number of adolescents mentioned boredom of homemade foods (18.8%) and curiosity (34.4%) as major reasons (Figure A of Supplementary File 2) for consumption which is familiar with other studies conducted in Nepal.^27^ This consistency might be the outcome of similar mindset of adolescents towards the processed foods. Level of knowledge was not found to be associated with the practice of sugary foods and drinks which is consistent with the findings of another study conducted in schools of Kathmandu by Paudel et al.^30^

Out of 768 adolescents, 6.38% of the respondents were found to have high BMI considerably categorized into overweight and obese. These results are consistent with those of Acharya et al and Khatri et al whose findings were around and less than ten percent.^37,38^ Overweight and Obesity among the adolescents was found to be significantly associated with consumption of sugary foods and beverages. The odds of overweight/obesity were higher among the adolescents consuming sugary foods and beverages compared to the ones not consuming. This finding is consistent with that of Singh et al. study done in adolescents among Kathmandu valley which showed a significantly higher risk of being overweight/obese: who consumed soft drink ≥1 time/day compared to < 1 time/day in past 1 months of the study (RRR = 5.44, 95% CI: 2.93– 10.10)^39^ Higher proportion of being overweight or obese was seen among adolescents who consumed more fast foods than the less frequent consumer was observed in the study done in Mangalore city of India as well.^33^ However, on the contrary, no association was found between KAP on SSB intake and body mass index among Malay adolescents in the study conducted by Teng et al.^36^ This conflicting study results could be associated with the nature of the study as our study didn’t consider factors such as time period of consumption.

## CONCLUSION

The findings revealed a good level of knowledge, moderate level of attitude, and poor level of lifestyle choices amongst sugary foods and beverages intake of the participants. Presence of adequate knowledge regarding sugary foods and beverages shows that the participants were aware regarding presence of higher amount of free sugar in sugary foods and beverages alongside their health effects most of them being aware about the weight gain as major consequences of the consumption of sugary foods and beverages. However, they were not found to be attentive and concerned towards reducing the intake of such food items as the consumption rate was really higher amongst study adolescents’ population. The food choices were still inclined towards the foods with higher amount of free sugar. Number of adolescents were found to be overweight and obese as well. Large proportion of the respondents were found to have normal weight. Also, consumption of the sugary foods and beverages was found to be associated with higher body weight among adolescents, where higher number of adolescents with more weight were consuming the sugary foods and beverages. Finally, interventions focused around habitual changes among adolescents are required to develop long-term behavior change strategies to reduce the consumption of sugary foods and beverages thus reducing the risk of overweight and obesity as well in order to improve the overall well-being of adolescents. The government of Nepal should strictly standardize and regulate consumption of sugar-rich foods and beverages among adolescents.

## Supporting information

Supplementary File 1: Tool

## DECLARATIONS

## Acknowledgement

We would to like to thank all school authorities and students who participated in the study. The authors acknowledge Office of the Municipal Office, Education Section and Health Section of Nagarjun Municipality for the coordination offered during the study. We are thankful to Department of Public Health, Manmohan Memorial Institute of Health Sciences for their immense support.

## Contributors

LG and MT contributed equally to this paper. LG and MT were responsible in conceptualizing the study, designing the protocol and development of research tools. MT was involved in collection of data and reviewing the literature. MT were involved in data curation alongside formal statistical analysis. PMS, PP, AB and MT were responsible for writing the original draft of the manuscript. LG and KP were responsible for critically reviewing and supervising the manuscript providing substantial input. MT is the guarantor of the manuscript. All authors read and approved the final manuscript.

## Ethics approval

Institutional Review Committee, Manmohan Memorial Institute of Health Sciences, Institute of Medicine, Tribhuvan University, Nepal

## Data availability statement

All data analyzed during this study are available from the corresponding author on reasonable request.

