## Supplementary File 1: Tool for "Cross sectional study on behavioral determinants associated with sugary foods and beverages and its corelates with body mass index among adolescents"

### SUPPLEMENTARY FILE 1: QUESTIONNAIRE (ENGLISH)

| S.N | Questions | Responses | | Skip pattern |
|---|---|---|---|---|
| **SECTION A- SOCIO DEMOGRAPHIC CHARACTERISTICS** | | | | |
| 1. | Age (in years) | | | |
| 2. | Sex | a. Male<br>b. Female | c. Others | |
| 3. | Ethnicity | a. Brahmin/ Chhetri<br>b. Janajati<br>c. Madhesi | d. Muslim<br>e. Dalit<br>f. Others | |
| 4. | Religion | a. Hindu | b. Other than Hindu | |
| 5. | Monthly income of family | Rs. | | |
| 6. | Who do you live with? | a. With parents<br>b. In hostel<br>c. With relatives<br>d. Others ………………………………….. | | |
| **SECTION B- PARENTAL CHARACTERISTICS** | | | | |
| 7. | Educational status of mother | a. Illiterate<br>b. Literate<br>c. Lower secondary level (6-8) | d. Secondary level (9-12)<br>e. Bachelors and above | |
| 8. | Occupation of mother | a. Agriculture<br>b. Labor<br>c. Job service<br>d. Business/Shop | e. Homemaker<br>f. Foreign employement<br>g. Others……………………………. | |
| 9. | Educational status of father | a. Illiterate<br>b. Literate<br>c. Lower secondary level (6-8) | d. Secondary level (9-12)<br>e. Bachelors and above | |
| 10. | Occupation of father | a. Agriculture<br>b. Labor<br>c. Job service<br>d. Business/Shop | e. Homemaker<br>f. Foreign employement<br>g. Others……………………………. | |
| **SECTION C- PERSONAL AND BEHAVIORAL CHARACTERISTICS** | | | | |
| 11. | Weight (to be taken by researcher) | | | (in kg) |
| 12. | Height (to be taken by researcher) | | | (in cm) |
| **SECTION D- KNOWLEDGE REGARDING SUGARY FOODS AND BEVERAGES AND ITS IMPACT ON HEALTH** | | | | |
| 13. | What is the meaning of sugary foods and drinks? | a. Foods and drinks with high amount of energies (calories)<br>b. Foods and drinks with adequate essential nutrients<br>c. None of them<br>d. Don't know | | |
| 14. | Are the sugary foods and beverages harmful for your health? | a. Yes they are harmful<br>b. No they are not harmful<br>c. Don't know | | |

| | What's your view on questions mentioned below? | (True) | (False) | (Don't know) |
|---|---|---|---|---|
| 15. | Foods and drinks with excess amount of sugar increases our weight and causes obesity. | | | |
| 16. | Foods and drinks with excess amount of sugar do not cause dental caries. | | | |
| 17. | Foods and drinks with excess amount of sugar causes stomach pain and bloating. | | | |
| 18. | Foods and drinks with excess amount of sugar causes various nutrtional deficiencies. | | | |
| 19. | Foods and drinks with excess amount of sugar increases the risk of causing high blood pressure and diabetes. | | | |

| | |
|---|---|
| 20 | Foods and drinks with excess amount of sugarmay cause stones in kidney. |
| 21. | Foods and drinks with excess amount of sugar do not cause any heart related diseases. |

**SECTION E- ATTITUDE TOWARDS SUGARY FOODS AND BEVERAGES**

| | **Statements** | **Categories** | | |
|---|---|---|---|---|
| | | **Agree** | **Neutral** | **Diasgree** |
| 22. | Sugary foods and drinks contain high amount of sugar content. | | | |
| 23. | Sugary foods and drinks contain harmful coloring agents to give artificial colour to them. | | | |
| 24. | Sugary foods and drinks contain various sweetening agents to enhance their taste. | | | |
| 25. | Sugary foods and drinks are very fresh foods. | | | |
| 26. | Sugary foods and drinks causes rapid weight gain. | | | |
| 27. | I feel that I should control sugary food and drinks consumption habit and decreases the intake of those. | | | |
| 28. | I will be healthy and free from diseases if I controlled the intake of sugary foods and drinks | | | |
| 29. | Excessive intake of sugary foods and drinks can increase my weight rapidly and make me overweight. | | | |
| 30. | My parents tell me not to eat or eat very less sugary foods and drinks. | | | |
| 31. | I can save my lots of money if I control myself from eating such sugary foods and drinks. | | | |
| 32. | I may be addicted to sugary foods and drinks if I didn't control my habit now. | | | |
| 33. | I like the taste and enjoy eating sugary foods and sugary beverages. | | | |
| 34. | But if asked, I can also stop eating and drinking such foods and drinks completely. | | | |

**SECTION F- PRACTICE REGARDING SUGARY FOODS AND BEVERAGES**

| | | | |
|---|---|---|---|
| 35. | Do you consume sugary foods and beverages? | a. Yes<br>b. No | |
| 36. | What are the sugary foods and drinks you consumed in the last week ? | | |
| 37. | What is the main source of information for sugary foods and drinks for you? | a. Social media<br>b. Advertisments<br>c. Friends<br>d. Siblings | e. Parents<br>f. Observation in stores<br>g. Others …………………… |
| 38. | Do you prefer sugary foods and beverages over usual meals as an alternative to regular meals? | a. Yes, I prefer<br>b. No, I don't prefer | |
| 39. | What are the major reasons for consumption of sugary foods and beverages? | a. Felt bored of home foods<br>b. Curiosity to try new foods<br>c. Favorite leisure time activity<br>d. Friends influence<br>e. Siblings influence | f. Advertisement influence<br>g. Quick to eat and finish<br>h. Affordable price<br>i. Good taste<br>j. (Others) …………………… |
| 40. | What is the most usual time to consume sugary foods and beverages? | a. Before school as breakfast<br>b. At school during lunch | c. After school<br>d. Whenever available |

Gautam L, Thapa M et al. (2023)
