## Supplementary File 2: Table and Figure for "Cross sectional study on behavioral determinants associated with sugary foods and beverages and its corelates with body mass index among adolescents"

### SUPPLEMENTARY FILE 2: TABLES AND FIGURE

**Table A.** Knowledge on sugary foods and beverages and their harmful effects

| Indicator category | Frequency | Percentage |
|---|---|---|
| Level of knowledge | | |
| Adequate knowledge | 646 | 84.1 |
| Inadequate knowledge | 122 | 15.9 |
| Meaning of sugary items | | |
| Contains high calories | 359 | 46.7 |
| Full of good nutrients | 53 | 6.9 |
| None of them | 114 | 14.9 |
| Don't know | 253 | 32.9 |
| Harmfulness of the items | | |
| Harmful | 625 | 81.4 |
| Not harmful | 39 | 5.1 |
| Don't know | 104 | 13.5 |
| Awareness of ill effects associated with consumption of sugary items | | |
| Weight gain & obesity | 655 | 87.2 |
| Dental caries | 688 | 89.6 |
| Digestive issues | 562 | 74.8 |
| Nutritional deficiency | 520 | 69.2 |
| High BP and diabetes | 657 | 87.5 |
| Risk of kidney stones | 341 | 45.4 |
| Heart related disease | 172 | 77.1 |

**Table B.** Attitude towards sugary foods and beverages

| Indicator category | Frequency | Percentage |
|---|---|---|
| Level of attitude | | |
| Positive attitude | 461 | 60.0 |
| Negative attitude | 307 | 40.0 |
| Perceived intake habit | | |
| Likes the taste and enjoys eating such foods | 449 | 58.5 |
| Neutral regarding the statement | 179 | 23.3 |
| Disagrees on enjoying eating such foods | 140 | 18.2 |
| Parents pressure against consumption | | |
| Agree | 602 | 78.4 |
| Neutral | 121 | 15.8 |
| Disagree | 45 | 5.9 |

**Table C.** Practice of sugary foods and beverages among respondents

| Indicator category | Frequency | Percentage |
|---|---|---|
| Consumption of sugary foods and beverages | | |
| Yes | 649 | 84.5 |
| No | 119 | 15.5 |
| Types of sugary foods and beverages consumed (*n=1222) | | |
| Processed sweet foods | 132 | 20.4 |
| Confectionary/ Bakery | 444 | 68.5 |
| Desert sweets | 137 | 21.1 |
| Sweet hot beverages | 79 | 12.2 |
| Cold beverages | 430 | 66.4 |
| Source of information (*n=804) | | |
| Social media | 182 | 29.0 |
| Advertisements | 175 | 27.9 |
| Friends | 124 | 19.8 |
| Siblings | 44 | 7.0 |
| Parents | 92 | 14.7 |
| Observation in stores | 187 | 29.8 |
| Preference as an alternative over usual meals (n=649) | | |
| Yes | 180 | 27.7 |
| No | 469 | 72.3 |
| Usual time for consumption (n=649) | | |
| As early breakfast | 39 | 6.0 |
| At school during lunch | 117 | 18.0 |
| After school | 230 | 35.5 |
| Whenever available | 263 | 40.5 |

**Table D.** Knowledge, Attitude and Practice of sugary foods and beverages among respondents

| Behavioral determinants | Frequency | Percentage |
|---|---|---|
| Knowledge | | |
| Adequate | 646 | 84.1 |
| Inadequate | 122 | 15.9 |
| Attitude | | |
| Positive attitude | 461 | 60.0 |
| Negative attitude | 307 | 40.0 |
| Practice (Consumption) | | |
| Yes | 649 | 84.5 |
| No | 119 | 15.5 |

**Table E:** Bivariate analysis of knowledge level and socio-demographic variables

| Demographic Characteristics | Level of knowledge | | χ2 | p-value |
|---|---|---|---|---|
| | Inadequate n (%) | Adequate n (%) | | |
| Type of school | | | | |
| Public | 55 (21.7) | 199 (78.3) | 9.45 | **<0.01*** |
| Private | 67 (13.0) | 447 (87.0) | | |
| Grade of study | | | | |
| Eight | 48 (18.8) | 207 (81.2) | 2.46 | 0.29 |
| Nine | 37 (14.5) | 218 (85.5) | | |
| Ten | 37 (14.3) | 221 (85.7) | | |
| Age (in years) | | | | |
| ≤ 14 | 49 (13.4) | 316 (86.6) | 3.15 | 0.07 |
| > 14 | 73 (18.1) | 330 (81.9) | | |
| Sex of the participant | | | | |
| Male | 60 (15.2) | 335 (84.8) | 0.29 | 0.58 |
| Female | 62 (16.6) | 311 (83.4) | | |
| Ethnicity | | | | |
| Brahmin/Chhetri | 51 (15.0) | 290 (85.0) | 0.58 | 0.74 |
| Janajati | 65 (16.9) | 320 (83.1) | | |
| Others | 6 (14.3) | 36 (85.7) | | |
| Religion | | | | |
| Hindu | 97 (16.7) | 485 (83.3) | 1.09 | 0.29 |
| Others | 25 (13.4) | 161 (86.6) | | |
| Monthly income of family | | | | |
| <15,000 NPR | 20 (27.0) | 54 (73.0) | 9.95 | **0.04*** |
| 15,001-30,000 NPR | 45 (17.4) | 214 (82.6) | | |
| 30,001-45,000 NPR | 23 (13.8) | 144 (86.2) | | |
| 45,001-60,000 NPR | 19 (13.1) | 126 (86.9) | | |
| >60,001 NPR | 15 (12.2) | 108 (87.8) | | |
| Living status of participants | | | | |
| With parents | 104 (14.9) | 596 (85.1) | 6.25 | **0.01*** |
| Others | 18 (26.5) | 50 (73.5) | | |
| **Parental demographics** | | | | |
| Educational status of mother | | | | |
| Illiterate | 26 (21.1) | 97 (78.9) | 3.29 | **<0.01*** |
| Literate | 21 (18.9) | 90 (81.1) | | |
| Primary level | 38 (20.2) | 150 (79.8) | | |
| Secondary level | 30 (11.4) | 234 (88.6) | | |
| Bachelors and above | 7 (8.5) | 75 (91.5) | | |
| Mother's occupation | | | | |
| Job service | 22 (17.3) | 105 (82.7) | 1.93 | 0.58 |
| Business/Shop | 22 (15.7) | 118 (84.3) | | |
| Homemaker | 46 (14.0) | 282 (86.0) | | |
| Others | 32 (18.5) | 141 (81.5) | | |
| Educational status of father | | | | |
| Illiterate | 9 (14.3) | 54 (85.7) | 8.61 | 0.07 |
| Literate | 18 (21.4) | 66 (78.6) | | |
| Primary level | 34 (20.1) | 135 (79.9) | | |

| | | | | |
|---|---|---|---|---|
| Secondary level | 48 (15.2) | 267 (84.8) | | |
| Bachelors and above | 13 (9.5) | 124 (90.5) | | |
| Father's occupation | | | | |
| Job service | 38 (15.0) | 215 (85.0) | | |
| Business/Shop | 31 (14.2) | 187 (85.8) | 8.04 | **0.04*** |
| Foreign employment | 15 (11.6) | 114 (88.4) | | |
| Others | 38 (22.6) | 130 (77.4) | | |

***p value <0.05**

**Table F:** Bivariate analysis of attitude and socio-demographic variables

| Demographic Characteristics | Level of attitude | | χ2 | p-value |
|---|---|---|---|---|
| | Negative n (%) | Positive n (%) | | |
| Type of school | | | | |
| Public | 103 (40.6) | 151 (59.4) | 0.05 | 0.81 |
| Private | 204 (39.7) | 310 (60.3) | | |
| Grade of study | | | | |
| Eight | 93 (36.5) | 162 (63.5) | 1.95 | 0.37 |
| Nine | 106 (41.6) | 149 (58.4) | | |
| Ten | 108 (41.9) | 150 (58.1) | | |
| Age (in years) | | | | |
| ≤ 14 | 139 (38.1) | 226 (61.9) | 1.03 | 0.30 |
| > 14 | 168 (41.7) | 235 (58.3) | | |
| Sex of the participant | | | | |
| Male | 184 (46.6) | 211 (53.4) | 14.80 | **<0.01*** |
| Female | 123 (33.0) | 250 (67.0) | | |
| Ethnicity | | | | |
| Brahmin/Chhetri | 143 (41.9) | 198 (58.1) | 1.12 | 0.56 |
| Janajati | 149 (38.7) | 236 (61.3) | | |
| Others | 15 (35.7) | 27 (64.3) | | |
| Religion | | | | |
| Hindu | 237 (40.7) | 345 (59.3) | 0.56 | 0.45 |
| Others | 70 (37.6) | 116 (62.4) | | |
| Monthly income of family | | | | |
| <15,000 NPR | 33 (44.6) | 41 (55.4) | 1.67 | 0.79 |
| 15,001-30,000 NPR | 102 (39.4) | 157 (60.6) | | |
| 30,001-45,000 NPR | 67 (40.1) | 100 (59.9) | | |
| 45,001-60,000 NPR | 53 (36.6) | 92 (63.4) | | |
| >60,001 NPR | 52 (42.3) | 71 (57.7) | | |
| Living status of participants | | | | |
| With parents | 278 (39.7) | 422 (60.3) | 0.22 | 0.63 |
| Others | 29 (42.6) | 39 (57.4) | | |
| **Parental demographics** | | | | |
| Educational status of mother | | | | |
| Illiterate | 45 (36.6) | 78 (63.4) | 11.27 | **0.02*** |
| Literate | 38 (34.2) | 73 (65.8) | | |
| Primary level | 80 (42.6) | 108 (57.4) | | |
| Secondary level | 121 (45.8) | 143 (54.2) | | |

| | | | | |
|---|---|---|---|---|
| Bachelors and above | 23 (28.0) | 59 (72.0) | | |
| Mother's occupation | | | | |
| Job service | 43 (33.9) | 84 (66.1) | | |
| Business/Shop | 57 (40.7) | 83 (59.3) | 3.51 | 0.31 |
| Homemaker | 130 (39.6) | 198 (60.4) | | |
| Others | 77 (44.5) | 96 (55.5) | | |
| Educational status of father | | | | |
| Illiterate | 23 (36.5) | 40 (63.5) | | |
| Literate | 30 (35.7) | 54 (64.3) | 1.40 | 0.84 |
| Primary level | 67 (39.6) | 102 (60.4) | | |
| Secondary level | 129 (41.0) | 186 (59.0) | | |
| Bachelors and above | 58 (42.3) | 79 (57.7) | | |
| Father's occupation | | | | |
| Job service | 90 (35.6) | 163 (64.4) | | |
| Business/Shop | 90 (41.3) | 128 (58.7) | 3.23 | 0.35 |
| Foreign employment | 54 (41.9) | 75 (58.1) | | |
| Others | 307 (40.0) | 95 (56.5) | | |

***p value <0.05**

**Table G:** Bivariate analysis of practice and socio-demographic variables

| **Demographic Characteristics** | **Consumption status** | | **χ2** | **p-value** |
|---|---|---|---|---|
| | **No n (%)** | **Yes n (%)** | | |
| Type of school | | | | |
| Public | 49 (19.3) | 205 (80.7) | 4.17 | **0.04*** |
| Private | 70 (13.6) | 444 (86.4) | | |
| Grade of study | | | | |
| Eight | 43 (16.9) | 212 (83.1) | | |
| Nine | 28 (11.0) | 227 (89.0) | 6.23 | **0.04*** |
| Ten | 48 (18.6) | 210 (81.4) | | |
| Age (in years) | | | | |
| ≤ 14 | 47 (12.9) | 318 (87.1) | 3.64 | 0.05 |
| > 14 | 72 (17.9) | 331 (82.1) | | |
| Sex of the participant | | | | |
| Male | 82 (20.8) | 313 (79.2) | 17.21 | **<0.01*** |
| Female | 37 (9.9) | 336 (90.1) | | |
| Ethnicity | | | | |
| Brahmin/Chhetri | 56 (16.4) | 285 (83.6) | | |
| Janajati | 57 (14.8) | 328 (85.2) | 0.41 | 0.81 |
| Others | 6 (14.3) | 36 (85.7) | | |
| Religion | | | | |
| Hindu | 93 (16.0) | 489 (84.0) | 0.43 | 0.51 |
| Others | 26 (14.0) | 160 (86.0) | | |
| Monthly income of family | | | | |
| <15,000 NPR | 12 (16.2) | 62 (83.8) | | |
| 15,001-30,000 NPR | 46 (17.8) | 213 (82.2) | 3.01 | 0.55 |
| 30,001-45,000 NPR | 27 (16.2) | 140 (83.8) | | |
| 45,001-60,000 NPR | 20 (13.8) | 125 (86.2) | | |

| | | | | |
|---|---|---|---|---|
| >60,001 NPR | 14 (11.4) | 109 (88.6) | | |
| Living status of participants | | | | |
| With parents | 110 (15.7) | 590 (84.3) | 0.29 | 0.58 |
| Others | 9 (13.2) | 59 (86.8) | | |
| **Parental demographics** | | | | |
| Educational status of mother | | | | |
| Illiterate | 23 (18.7) | 100 (81.3) | 6.61 | 0.15 |
| Literate | 17 (15.3) | 94 (84.7) | | |
| Primary level | 36 (19.1) | 152 (80.9) | | |
| Secondary level | 36 (13.6) | 228 (86.4) | | |
| Bachelors and above | 7 (8.5) | 75 (91.5) | | |
| Mother's occupation | | | | |
| Job service | 26 (20.5) | 101 (79.5) | 9.89 | **0.01*** |
| Business/Shop | 23 (16.4) | 117 (83.6) | | |
| Homemaker | 36 (11.0) | 292 (89.0) | | |
| Others | 34 (19.7) | 139 (80.3) | | |
| Educational status of father | | | | |
| Illiterate | 13 (20.6) | 50 (79.4) | 6.74 | 0.15 |
| Literate | 14 (16.7) | 70 (83.3) | | |
| Primary level | 32 (18.9) | 137 (81.1) | | |
| Secondary level | 47 (14.9) | 268 (85.1) | | |
| Bachelors and above | 13 (9.5) | 124 (90.5) | | |
| Father's occupation | | | | |
| Job service | 45 (17.8) | 208 (82.2) | 3.95 | 0.26 |
| Business/Shop | 34 (15.6) | 184 (84.4) | | |
| Foreign employment | 13 (10.1) | 116 (89.9) | | |
| Others | 27 (16.1) | 141 (83.9) | | |

***p value <0.05**

**Table H.** Body Mass Index (BMI) of the respondents

| Categories: Range of BMI | Frequency | Percentage |
|---|---|---|
| Underweight | 332 | 43.2 |
| Normal | 387 | 50.4 |
| Overweight and Obese | 49 | 6.4 |

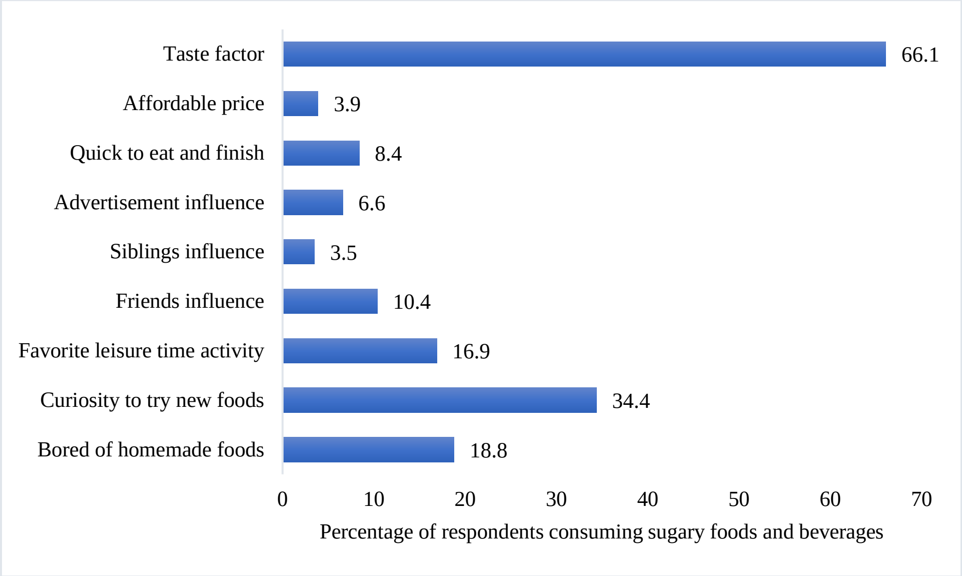


**Figure A.** Reasons for consumption of sugary foods and beverages among respondents
